# Effects of Subthalamic Deep Brain Stimulation on Emotion Processing in Parkinson’s Disease

**DOI:** 10.1101/2025.08.25.25333958

**Authors:** Dilara Bingöl, Clara Klimaschewski, Lars Timmermann, David Pedrosa, Josefine Waldthaler

**Author notes:** contributed equally to this work (David Pedrosa, Josefine Waldthaler). **Correspondence to:** Josefine Waldthaler,; Present Address: Department of Clinical Neuroscience, Karolinska Institutet, Nobels väg 9, D2, 171 77 Stockholm.

## Abstract

**Background:** Processing of emotion plays a critical role in social interaction. People with Parkinson’s Disease (PwPD) may experience impaired facial emotion recognition for negative emotions. Previous evidence regarding the effects of deep brain stimulation in the subthalamic nucleus (STN-DBS) is inconclusive.

**Objective:** To investigate the impact of STN-DBS on emotion processing in PwPD controlling for effects of disease progression and DBS-induced changes in cognitive ability. The secondary aim was to explore the effect of post-DBS adjustments of dopamine replacement therapy.

**Methods:** 32 cognitively intact PwPD (16 DBS, 16 non-DBS) and 22 healthy controls performed tasks assessing recognition and discrimination of emotions and an emotional Go/No-Go paradigm, complemented by task versions using non-emotional stimuli. Baseline assessments were conducted two months before DBS surgery and followed up three months postoperatively. Mixed-effects models analysed group-by-time interactions on task performance measured as accuracy and reaction time. Associations of longitudinal changes in non-motor symptoms and medication dosages with task performance were assessed using bootstrapped Pearson’s correlations.

**Results:** A significant DBS-associated increase in reaction times was found specifically in the emotional conditions, while there was no evidence for a DBS-effect on the accuracy of emotion recognition or discrimination. Post-DBS dose reduction of dopamine agonists was associated with improvement in emotion recognition.

**Conclusion:** STN-DBS does not lead to a generalized decline in emotional processing when controlling for important confounders. Instead, changes in response speed suggest altered processing dynamics. Adjustments in pharmacological treatment regimens contribute to altered FER after DBS surgery.

## 1. Introduction

Facial emotion recognition (FER) is crucial for understanding emotions and navigating social interactions. Our ability to decode others’ expressions allows us to interpret and respond to their emotional states. Impaired FER makes nonverbal communication challenging, potentially straining social relationships, and reducing quality of life [1].

In Parkinson’s disease (PD), FER deficits have been observed, particularly concerning negative emotions [2,3]. While some studies report generalized FER impairment [4], others support larger variability based on specific emotions, individual differences, or methodological factors [5,6]. Impairments in recognizing fear and disgust are among the most frequently reported deficits in PD [7,8].

When it comes to treatment of people with Parkinson’s Disease (PwPD), Deep Brain Stimulation in the subthalamic nucleus (STN-DBS) has been demonstrated to effectively alleviate motor symptoms and improve quality of life [9,10]. However, its effects on non-motor symptoms, including FER, remain less well understood, with previous studies yielding mixed results. Some research indicates a worsening of FER after STN-DBS, particularly for negative emotions [11–14]. In contrast, Duits and colleagues reported no significant effects on FER one year after surgery in the largest cohort to date, suggesting that initial deficits may be transient or related to the surgery itself rather than chronic DBS [15]. A differential impact of the so-called lesion effect has first been confirmed by Aiello and colleagues [12]. Another possible confounder are influences of dopamine replacement therapy. In addition to the effects levodopa and dopamine agonists themselves may exert on emotion processing, previous work suggests complex interactions with DBS [11].

Most previous studies lack non-emotional control tasks, which are essential for distinguishing deficits specific for emotional processing from broader more general DBS-associated cognitive alterations. Additionally, most previous research has primarily focused on accuracy, with comparatively little attention paid to reaction times and DBS-associated changes in response inhibition – both of which are critical for a comprehensive understanding of emotional processing [14,16]. Increased impulsivity – frequently reported as a side effect of both pharmacological treatment with dopamine agonists and STN-DBS [17] – may interfere with the ability to interpret and respond appropriately to emotional cues [16,18].

In this study, we addressed several limitations of previous work by incorporating well-matched control groups and non-emotional control conditions in a within-participant longitudinal design in off-medication state. This allows us to differentiate between long-term DBS-related emotional disturbances and those attributable to general disease progression and short-lasting effects of levodopa or DBS surgery itself. In addition, we aimed to explore the effects of post-DBS adjustments of dopamine replacement therapy with a particular interest on the potential long-lasting effects of dopamine agonists.

## 2. Materials and methods

### 2.1. Ethical approval

The study was approved by the local Ethics Committee at University Hospital Marburg (reference: 118/21) and adhered to the principles described in the Declaration of Helsinki. Prior to their inclusion, all participants provided written informed consent.

### 2.2. Participants

A total of 46 cognitively intact PwPD diagnosed based on Movement Disorder Society (MDS) diagnostic criteria [19], and 32 healthy control individuals (HC) were recruited from the Department of Neurology at the University Hospital of Marburg between February 2022 and June 2023.

The PD cohort was divided into two groups based on whether they were scheduled for DBS surgery or not (non-DBS). Each participant underwent an initial baseline (BL) assessment, followed by a subsequent follow-up (FU) at least three months later (Figure 1). For individuals in the DBS group, BL assessments were conducted prior to surgery, while FU occurred three months after surgery. Surgical procedures, including preoperative stereotactical planning to target the dorsolateral aspect of the STN were conducted in accordance with local protocols for clinical care [20]. Correct lead placement in the dorsolateral portion of the STN was confirmed via postoperative CT scans.

**Figure 1:**
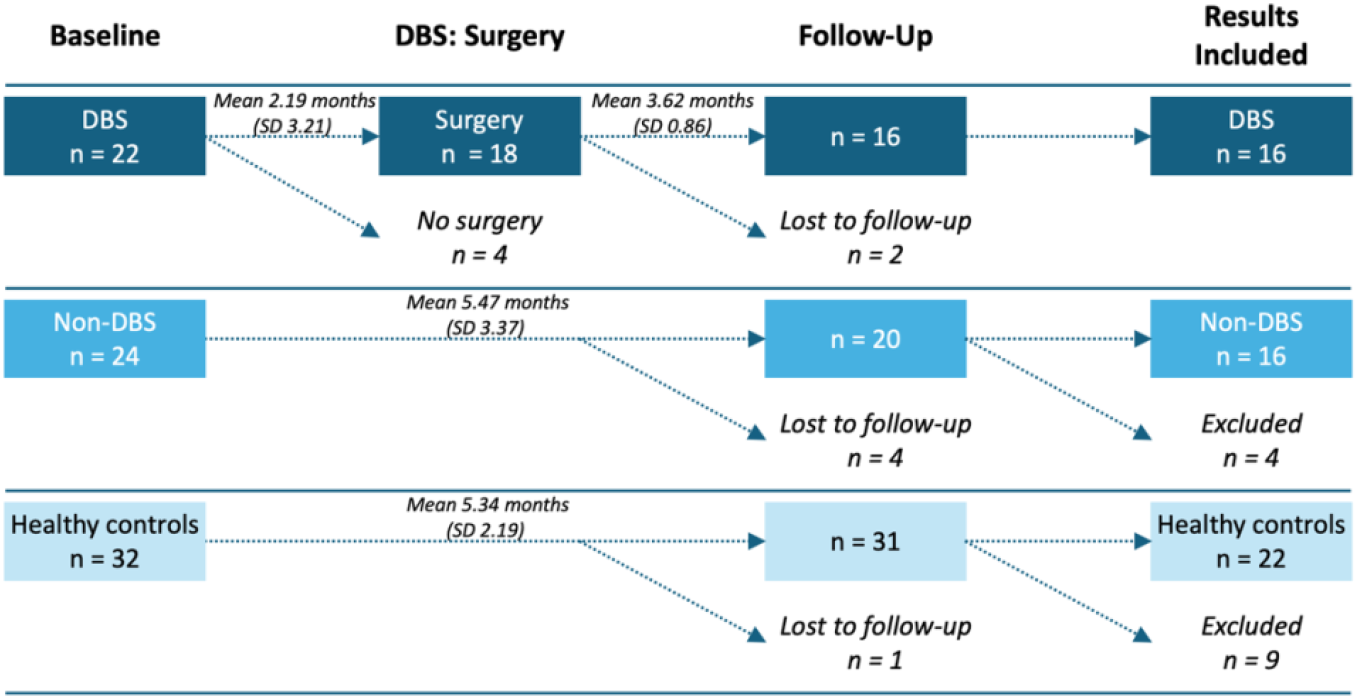
Overview of patient recruitment procedures and timeline.

Motor impairment was assessed using the Unified Parkinson’s Disease Rating Scale Part III (MDS-UPDRS-3) [21]. Levodopa Equivalent Daily Dose (LEDD) and dopamine agonist LEDD (DA-LEDD) were calculated according to [22].

Exclusion criteria, recruitment and matching procedures are provided in the Supplementary Material.

### 2.3. Cognitive assessment and Questionnaires

Cognition was assessed with the Montreal Cognitive Assessment (MoCA) [23] and the Frontal Assessment Battery (FAB) [24]. The PD groups completed the cognitive assessment in on-medication state after intake of their regular dose of dopamine replacement therapy.

Behavioral symptoms and quality of life related to PD were evaluated at baseline and follow-up using the Apathy Evaluation Scale (AES) [25], Beck’s Depression Inventory (BDI-II) [26], Hypomania Checklist-32 (HCL-32) [27], Questionnaire for Impulsive-Compulsive Disorders in Parkinson’s Disease (QUIP-RS) [28] and the 39-item Parkinson’s Disease Questionnaire (PDQ-39) [29].

### 2.4. Behavioral tasks

Three computer-based behavioral tasks were developed using custom-code in OpenSesame (Version: 3.3.11) [30] assessing response inhibition in an emotional Go/No-Go task, emotion discrimination and FER, each complemented by non-emotional control versions using neutral stimuli. In all emotional task conditions, static visual stimuli displaying emotional facial expressions from the ‘straight’ frontal perspective from the Karolinska Directed Emotional Faces (KDEF) database were used [31]. The identities and gender of the emotional face varied across trials to increase ecological validity.

All PwPD completed tasks after at least 12-hours withdrawal from all dopaminergic medication (“off-medication”), while those with STN-DBS performed follow-up tasks off-medication but with ongoing stimulation.

Each task began with at least three example trials to ensure understanding. Accuracy (proportion of correct responses) and response time (RT) were measured in the Go/No-Go and emotion discrimination tasks, while FER was assessed by accuracy alone.

#### 2.5.1. Emotional Go/No-Go task

Response inhibition was assessed with separate blocks of an emotional and non-emotional version of a Go/No-Go task with a 70/30 ratio. After a fixation cross with a random interstimulus interval between 250 and 750 ms, the trial began with a static visual stimulus consisting of a face displaying either a happy or angry emotional expression or a non-emotional character (X or K) (Figure 2). Upon the “go” stimulus, participants pressed the spacebar on a keyboard, while they were asked to withhold the response following the “no go” stimulus. The respective stimulus associated with the “go” and “no go” instruction switched between blocks. Four blocks were presented: two blocks with emotional stimuli interleaved with two blocks presenting neutral stimuli, resulting in a total of 100 trials per condition.

**Figure 2.**
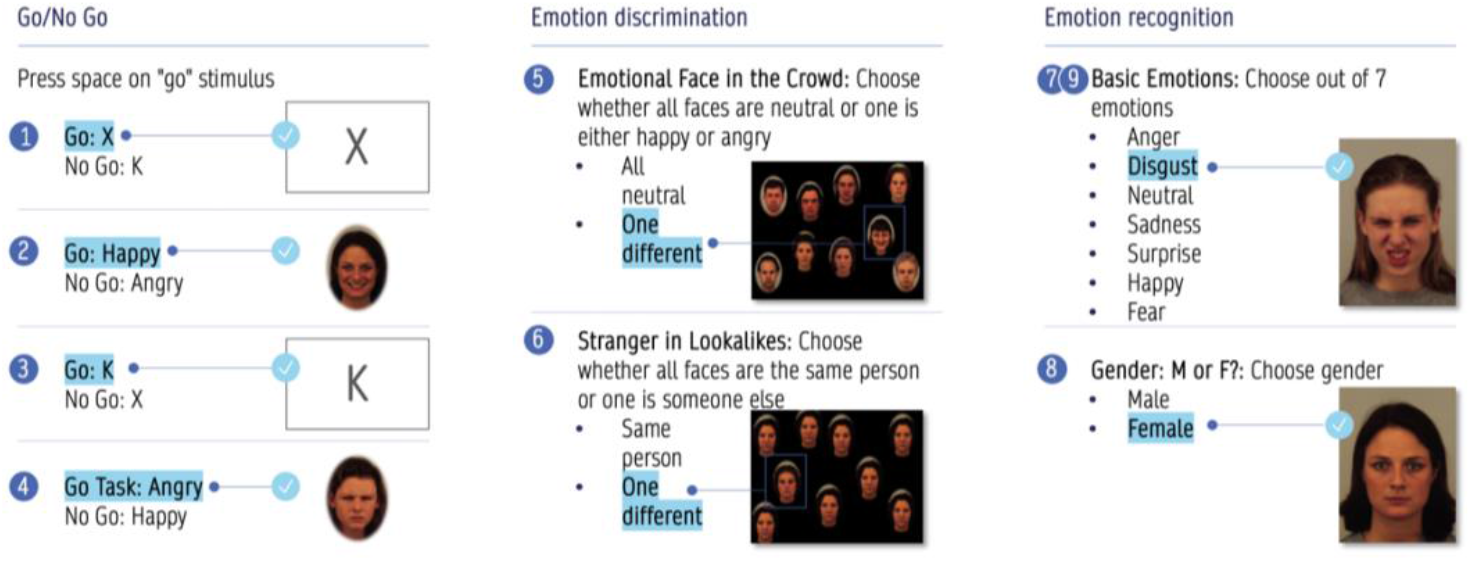
Task design.

#### 2.5.2. Emotion discrimination tasks

To assess participants’ ability to discriminate between two simultaneously presented emotions, we used a custom version of the “Face-in-the-crowd” task [32]. Each stimulus consisted of a randomly arranged array of nine different human faces (equally distributed across male and female). To reduce predictability, trials were designed without a grid pattern, preventing participants from anticipating the location of stimuli. Additionally, to prevent “central fixation bias” [33], the distractor was intentionally not placed in the centreer. The emotional version of the task included 60 pseudo-randomized stimuli, with 40 showing one emotional (20 happy and 20 angry) facial expression (the target) among eight neutral faces (the distractors), and 20 displaying nine neutral faces without a target. Participants pressed the left arrow key upon identifying a target or the right arrow key when they felt confident that all faces displayed the same expression. The non-emotional version of the task consisted of 42 trials. In 21 of these, the distractor was a “stranger” among a crowd of eight “look-a-likes” with the same identity, while the other half were arrays of nine “look-a-likes” without any distractor.

#### 2.5.3. Emotion recognition

The emotional version of the FER task required participants to identify the correct emotion displayed on a centrally presented face, while the non-emotional task involved identifying the perceived gender of the face. After they were presented the stimulus for 1000 ms, participants chose an emotion from a list of seven basic emotions: fear, sadness, disgust, happiness, surprise, anger, or neutral. After selecting an answer using the touch pad, they confirmed their choice by clicking the ‘ok’ button. Two emotional and one non-emotional pseudo-randomized block of this task were performed. The two emotional blocks included a total of 154 stimuli with 22 stimuli per emotion. Forty female and 37 male faces were presented in the non-emotional condition.

### 2.6. Data analysis

We utilized R programming language to conduct the analyses [34]. Trials with RT lower than 100 ms and above the individual mean plus three standard deviations were discarded. Absolute change scores between follow-up and baseline measurements were calculated for all outcome measures.

To assess for DBS effects on accuracy, we built separate logistic-mixed models for each of the three tasks using the lme4 package [35] with trial outcome (correct/error) as the dependent variable and group (PD/DBS/HC), condition (emotional/non-emotional version), time point (baseline/follow-up) and their interactions as fixed effects, adding each participant as random effect. As we hypothesized that change from baseline would depend on both group and condition, the three-way interaction term between all fixed effects was defined as the primary outcome of interest. The emmeans package was used to test the interaction contrasts for all effects in the model and to compile them into type-III-ANOVA-like results. In the Go/No-Go-task, commission and omission errors were analyzedanalysed separately. Regarding RT, we ran linear-mixed models similar to the logistic models defined for accuracy.

In a secondary analysis of each emotion in the FER task, we excluded the non-emotional condition, i.e., only modelling effects of group and time point, and their interactions.

Pearson correlations were used to explore association of changes in task performance with changes in total LEDD and dopamine agonist LEDD within the DBS group with 95% confidence intervals estimated using bootstrap resampling with 1,000 iterations.

## 3. Results

### 3.1. Participants’ clinical characteristics

The DBS group (n = 16, 3 female) had a mean age of 60.3 years (SD = 9.4) and a disease duration of 7.5 years (SD = 4.2). Baseline LEDD of 900.97 mg/day (SD = 313.36) decreased to 625.19 mg/day (SD = 268.59) after DBS surgery (p = 0.002, T = 2.13). Motor impairment, assessed by the MDS-UPDRS-III in the off-medication-state, showed a notable reduction from a baseline of 37.56 (SD = 11.59) to 25.30 (SD = 11.55) postoperatively (p = 0.01, T = 2.12 [pre- and postoperative value available in N = 10]).

In addition to matched age and sex across all three groups (HC: n = 22, 6 female, mean age 57.2 years (SD = 11.6), F = 0.42, p = 0.7), the non-DBS group (n = 16, 4 female, mean age 59.4 years, SD = 11.6) was matched for disease duration (mean 5.31 years, SD = 4.61, t = 1.41, p = 0.2) and Hoehn & Yahr stage (median = 2). In the non-DBS group, baseline LEDD was 536.22 mg/day (SD = 408.00), which slightly increased to 599.58 mg/day (SD = 430.74) at follow-up (p = 0.03, T = 2.13).

No significant changes in any of the scores assessing depression, hypomania, cognition and quality of life were found in any of the groups from baseline to follow-up

### 3.2. Behavioural tasks

#### 3.2.1. Emotional recognition

There was no significant three-way interaction effect of group, condition, and time point on overall emotion recognition accuracy including all emotions (F = 1.754, p = 0.173) (Figure 3).

**Figure 3:**
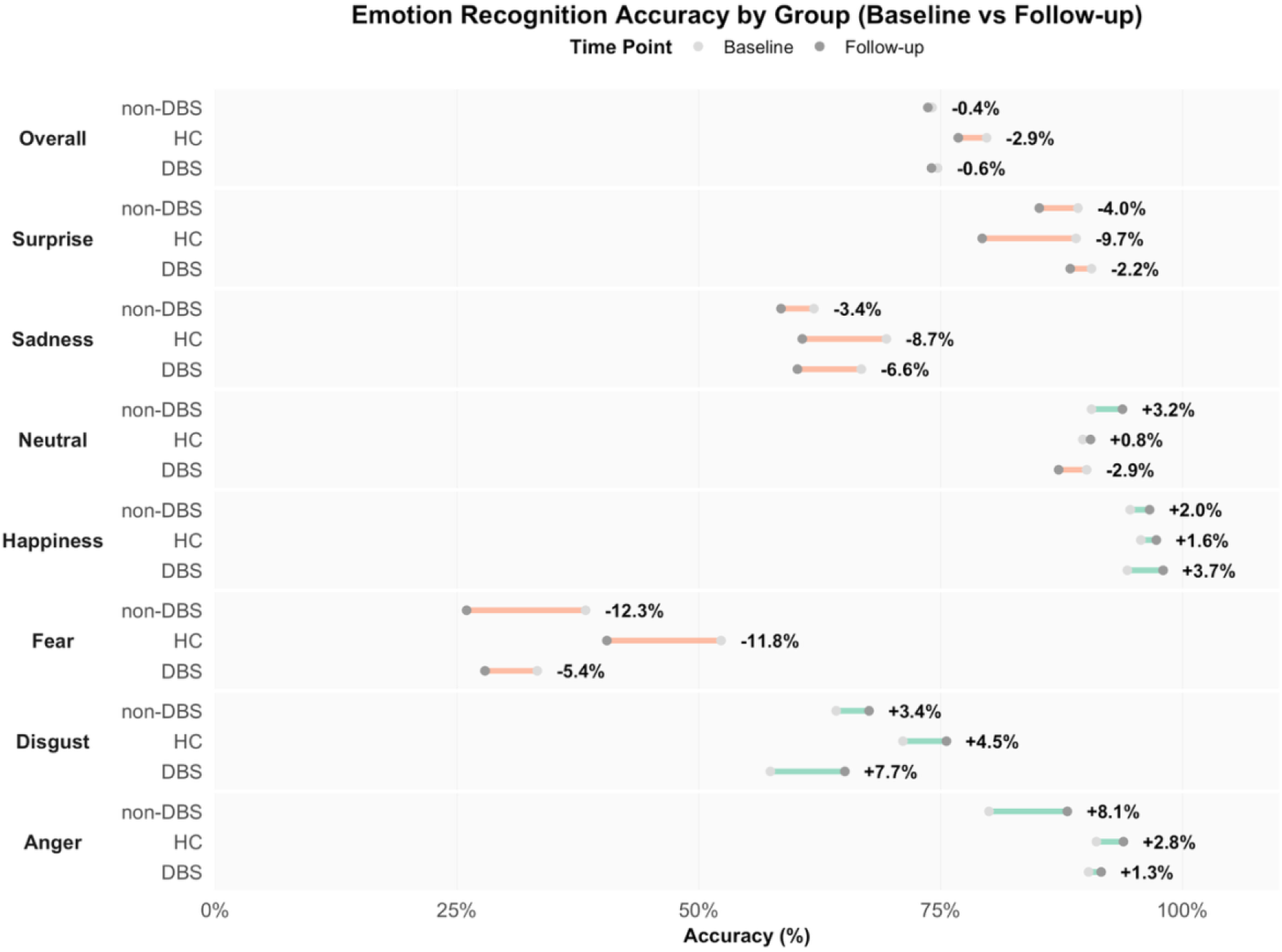
Mean accuracy of FER for all emotions by group and time point: Coloured lines connect baseline and follow-up accuracy values for each group: Orange lines indicate a decline in performance; green lines indicate improvement. Grey dots represent mean accuracy at each time point (light grey for baseline, dark grey for follow-up).

Further, there was no evidence for two-way interactions of group and time point in the secondary analysis for each emotion (all p >.05) (Supplementary Table 3). Notably, significant group effects in the models assessing disgust (F = 3.064, p = 0.047) and fear (F = 5.701, p = 0.003) became evident with both PD groups, DBS and non-DBS, performing worse than HC, regardless of time point.

#### 3.2.2. Emotion discrimination (Face in the crowd task)

There was no significant three-way interaction for group, condition and time point on accuracy (z = −0.154, p = 0.878 for HC vs. DBS, z = −1.167, p = 0.243 for non-DBS vs. DBS). However, a significant three-way interaction was observed for RT (z = 3.478, p < 0.001). Post-hoc comparisons by group and condition revealed a significant main effect of time point on RT in the DBS group during the emotional condition (F = 13.513, p < 0.001; increased RT at follow-up), as well as in the non-DBS group during the non-emotional condition (F = 27.156, p < 0.001; increased RT at follow-up) (Figure 4). No significant effects were observed for any other group/condition combinations (Supplementary table 4).

**Figure 4:**
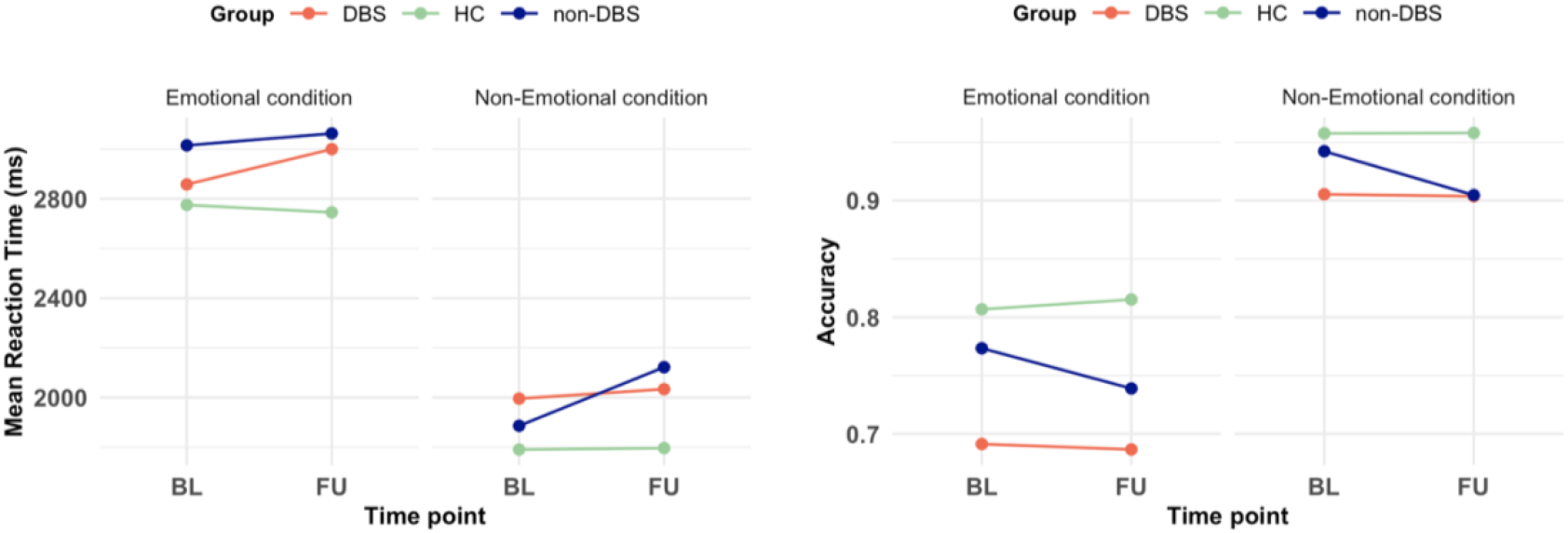
Reaction time and accuracy in the face in the crowd task: emotional vs. non-emotional conditions over time points: Lines represent each group over time: DBS (red), non-DBS (blue) and HC (green). “Emotional condition” includes trials presenting emotional stimuli (happy and angry combined).

Based on previous evidence suggesting either “happy” or “anger” superiority effects in the face-in-the-crowd-task, we next compared happy and angry distractors separately. There was a strong effect of distractor type evident in all groups with shorter RT (t = −7.317, p <0.01) and higher accuracy for happy distractors (z = 9.619, p = <0.01), i.e., a so-called happy superiority effect independent of group and time point.

#### 3.2.3. Emotional Go/No-Go task

There were no significant three-way interaction effects of group, condition, and time point for neither omission (F = 1.776, p = 0.169) nor commission error rates (F = 1.695, p = 0.184). However, we found a significant three-way interaction effect on RT (F = 7.179, p < 0.001). Post-hoc comparisons revealed that DBS-patients exhibited a significant RT increase exclusively in the emotional condition compared with non-DBS participants (p = 0.029) but not with HC (p = 0.152). Accuracy and RT in all conditions and groups are presented in Supplementary Table 5.

#### 3.2.4. Analyses including the DBS-group only

To compare with previous studies lacking control groups and non-emotional control conditions, we ran additional models including only the DBS group with time point as the independent effect and subject as random effect. Here, no evidence for a significant main effect of time point was found for overall emotion recognition (F = 0.344, p = 0.557). By emotion, recognition of disgust (F = 4.787, p = 0.029) and happiness (F = 6.036, p = 0.014) improved after STN-DBS surgery, whereas recognition of sadness (F = 3.689, p = 0.055) and fear (F = 2.894, p = 0.089) showed potential decline without reaching statistical significance.

In the face-in-the-crowd task, RT significantly increased over time (F = 13.513, p = < 0.01, while accuracy remained unchanged (F = 0.047, p = 0.829) mirroring our primary analysis.

In the emotional Go-No-Go task, commission errors (i.e., false-alarm responses during no-go trials) significantly decreased at follow-up (F = 4.861, p= 0.028), while omission errors remained unchanged (F = 0.385, p = 0.535). Response time also increased (F = 11.505 p < 0.001). Together, these results suggest improved response inhibition and reduced impulsivity in response to emotional stimuli with DBS.

#### 3.2.5. Association with postoperative adjustments of dopamine replacement therapy

As LEDD was reduced substantially in most patients after DBS surgery, we aimed to explore the effects of the adjustments of medication on our results. While all patients were tested in the practical “off” medication state, long-lasting effects of dopamine replacement therapy are not washed out entirely after 12-hour withdrawal. This is particularly true for dopamine agonists which also are known to impact affective and cognitive processing [36].

For the composite of all three emotional conditions, a statistically significant negative correlation was observed between LEDD change and accuracy change (r=−0.481, 95% CI [−0.78, −0.12]). No significant correlations were found for non-emotional conditions (r = −0.311, 95% CI [−0.76, 0.32]) or individual emotional tasks (Figure 5).

**Figure 5:**
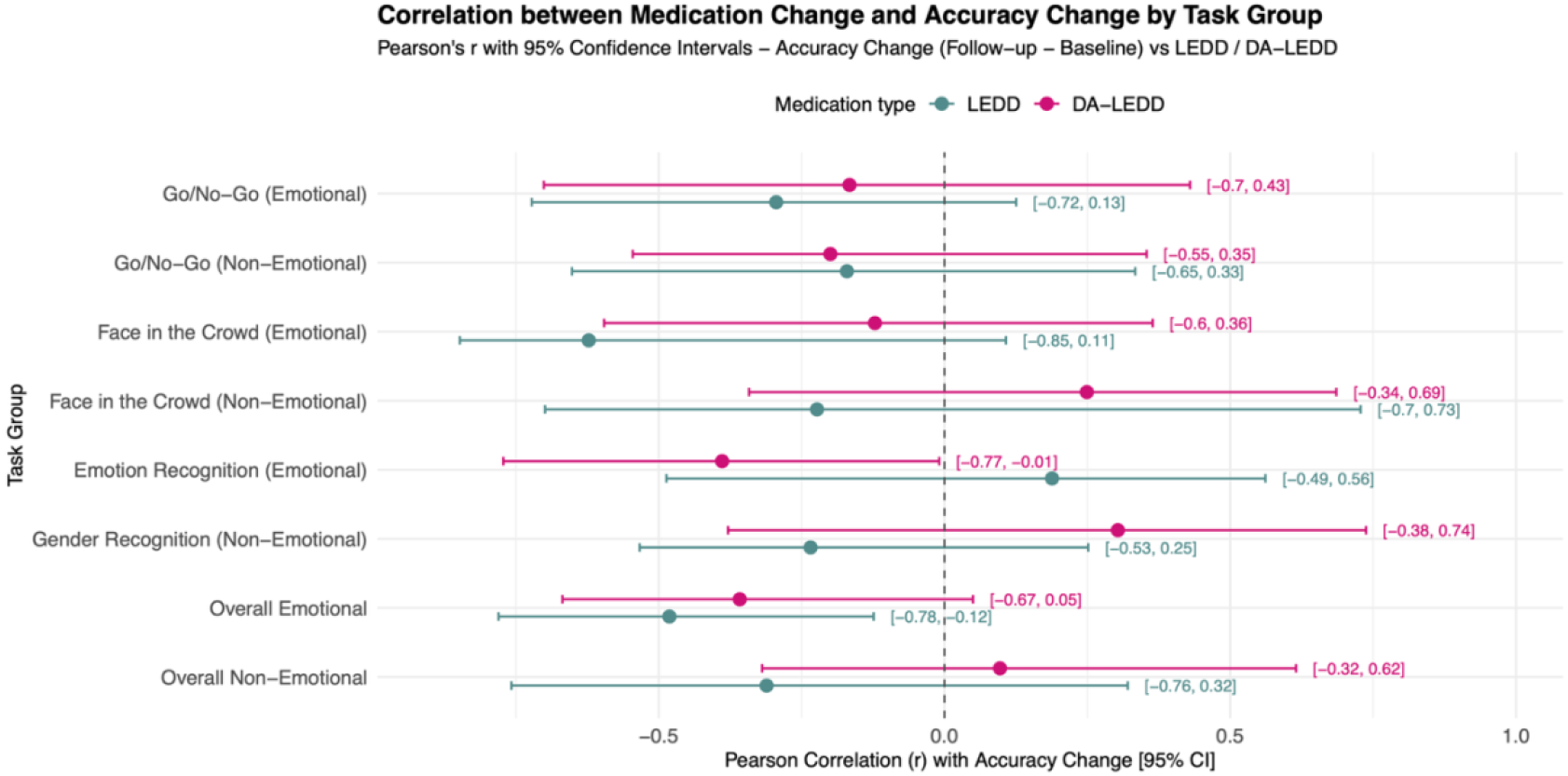
Correlation between medication change and accuracy change by condition by medication type (LEDD in blue and DA-LEDD in pink): Each dot represents the Pearson correlation coefficient (r) between absolute change in accuracy (follow-up minus baseline) and medication change (LEDD or DA-LEDD). Horizontal lines show 95% confidence intervals, estimated via 1000 bootstrap samples. The dashed vertical line at r = 0 indicates no association. Dots further to the right (r>0) indicate that medication reduction was associated with a performance decline. Dots further to the left (r<0) indicate that medication reduction was associated with improved performance. The “Overall Emotional” row aggregates accuracy across all emotional conditions, while the “Overall Non-Emotional” row aggregates accuracy across all non-emotional conditions.

A significant negative correlation was observed between dopamine agonist LEDD change (DA-LEDD), and FER accuracy change (r = −0.389, 95% CI [−0.77, −0.01]). No significant correlations were observed between LEDD or DA-LEDD changes and reaction time changes across conditions (Supplementary Figure 1).

## 4. Discussion

Emotional processing is a core component of human functioning, shaping our social interactions, our decision-making, and overall quality of life. While emotional symptoms are well-documented in PD, the influence of medical and surgical treatments - both of which may potentially alter emotional processing - remains an area of active investigation. While some studies reported impairments, particularly for negative emotions such as fear, sadness [7,37,38] or disgust [6,12] with DBS, others found no such effects [15]. Our findings support the latter as we found no decline of FER or emotion discrimination following STN-DBS compared with best medical treatment. Consistent with previous studies, PwPD exhibited deficits in FER for disgust and fear when compared with healthy individuals, regardless of treatment type [39].

Unexpectedly, participants with STN-DBS showed an increase in response times in the emotional Go/No-Go paradigm at follow-up compared with the non-DBS PD group, though their performance did not differ significantly from that of healthy controls. These findings suggest reduced impulsivity in the DBS cohort, contrasting with studies reporting disinhibition after STN-DBS [40,41]. One possible explanation is that DBS may enhance inhibitory control in emotionally salient contexts, potentially through modulation of prefrontal-subcortical circuits involved in cognitive control [42].

The significant DBS-induced increase in RT in the discrimination task that was limited to the emotional condition suggests that DBS differentially affects emotional versus non-emotional face processing speed. These findings align with evidence that emotional and non-emotional facial expressions are processed through distinct mechanisms [43]. The observation that RT, but not accuracy, were affected is consistent with findings that dopaminergic modulation can influence processing speed and cognitive effort without necessarily impacting task performance outcomes [44]. Alternatively, the RT increase might reflect a mechanism to compensate for a slightly reduced ability to discriminate emotional expressions that does not reach an extend where it becomes evident through decreasing accuracy. Interestingly, the non-DBS group also exhibited increased reaction times over time, but only in the non-emotional condition. Since this effect was absent in the DBS and HC groups, it is unlikely to stem from repeated testing alone where increased task familiarity could lead to more cautious responses. Instead, this may reflect cognitive slowing, possibly related to disease progression.

Regardless of DBS status, we observed a so-called happy superiority effect in the emotion discrimination task, with participants being significantly better and faster detecting happy compared to angry target emotions. This effect is consistent with previous research [45]. However, it is important to note that other studies have demonstrated an anger superiority effect [46]. These mixed findings suggest that both positive and negative facial expressions can confer a processing advantage, depending on methodological factors such as stimulus material, task demands and possibly participant age or context. Thus, while our results support a happy superiority effect in this sample and paradigm, further research is needed to clarify under which conditions each effect emerges and what mechanisms underlie these patterns in PD and with DBS treatment.

Given the known influence of dopaminergic modulation in emotional processing, we examined how postoperative changes in dopaminergic medication relate to performance changes. In the DBS group, larger post-DBS reductions in dopamine agonists were linked to improved accuracy in emotion recognition. These findings align with prior work showing that dopaminergic medication - particularly dopamine agonists – can exert domain-specific negative effects on cognition and behavior in PwPD [47]. Emotional processing appears especially sensitive to these dopaminergic changes, whereas non-emotional cognitive domains and processing speed remained unaltered in our sample. This underscores the need to account for both baseline medication and dose changes when interpreting post-DBS non-motor outcomes. Medication adjustments may mask or offset stimulation-related effects - for instance, cognitive improvement following agonist reduction could obscure subtle DBS-induced impairment of emotion processing. Future studies should include larger samples and statistical controls for medication status and dose changes to disentangle these interacting effects.

To contextualize our findings within the existing literature, we conducted an additional analysis focusing solely on the DBS group. This approach mirrors earlier research that assessed within-group changes in the absence of a non-DBS control group. In this analysis, an improvement in recognizing disgust and happiness was observed, alongside a trend toward reduced recognition of sadness and fear. This pattern partially aligns with previous research, in which sadness and fear impairments have been frequently reported post-DBS [14,38]. However, the unexpected improvement in disgust recognition is notable, as prior studies have more commonly documented decline in disgust recognition [11,12,14]. Our results support that these discrepancies may largely be explained by the lack of appropriate control groups in earlier studies. Alternatively, our null findings for negative emotions might reflect statistical power limitations rather than a true absence of effect. While our sample is relatively large for a DBS study, it remains smaller than some previous works that used larger datasets or metanalytical approaches [15,48]. Further, variability in outcomes could reflect differences in stimulation parameters or lead locations. It has been posited that dorsally placed leads may better preserve cognitive control, whereas ventral may disrupt limbic circuits [49]. In our sample, location of the DBS leads in the targeted dorsolateral aspect of the STN was confirmed postoperatively, and thus, our findings may not be transferable to leads in other locations within the STN or its surroundings.

Some additional limitations warrant consideration when interpreting our results. First, the study addressed a single postoperative time point, limiting the ability to evaluate potential long-term adaptation mechanisms. While prior studies with one-year follow-up reported no significant changes in emotion recognition after STN-DBS [15,50], suggesting that major effects are unlikely to emerge beyond the first year, this does not preclude the possibility of gradual or delayed changes. Longitudinal assessments across multiple time points would be valuable in determining whether previously observed post-DBS effects are transient or part of a broader adaptive trajectory.

An alternative explanation for the increased reaction times observed at follow-up in our DBS cohort is that they may reflect broader cognitive processing differences rather than effects specific to emotional stimuli. Tasks such as letter discrimination (e.g., “X” vs. “K”), are inherently simpler than interpreting facial expressions. Thus, differences in task complexity rather than emotional content alone may underlie the observed effects. Future research should include non-emotional control tasks with matched cognitive demands to further distinguish emotion-specific from general cognitive effects.

Finally, it is important to consider the potential long-lasting effects of dopaminergic medications. Although we ensured that all participants were assessed in a practically defined “off” state, residual effects—particularly from long-acting dopamine agonists—may have influenced the results as indicated by the identified association of dopamine agonist reduction and post-DBS changes in FER.

In summary, our findings indicate that STN-DBS does not lead to a generalized decline in emotional processing in PD when controlling for test-retest effects and disease progression with well-matched control groups. However, we observed significant increases in reaction time for DBS patients compared to non-DBS patients on both the Face-in-the-Crowd and emotional Go/No-Go tasks at follow-up, despite stable accuracy. This suggests that STN-DBS may subtly affect the efficiency rather than the accuracy of emotional stimulus processing. Notably, the slowing was not evident when DBS patients were compared to healthy controls, raising questions about compensatory mechanisms or the sensitivity of these tasks to disease-specific patterns.

To conclude, these results underscore the importance of using robust control designs and multiple behavioral indices to disentangle nuanced effects of STN-DBS on emotional function. Future studies should incorporate larger samples, multiple postoperative time points and imaging-based measures of DBS lead placement and connectivity to further elucidate how STN-DBS modulates emotional and cognitive circuits in PD.

## Data Availability

All data produced in the present study are available upon reasonable request to the authors.

## Acknowledgments

We thank all participants for their support, patience and cooperation. DB is grateful for the support from the Foundation of German Business (Stiftung der Deutschen Wirtschaft) – Klaus Murmann Fellowship, a scholarship awarded during medical studies, funded by the German Federal Ministry of Education and Research (BMBF).

## Author Contributions

**DB**: Investigation, Data curation, Software, Formal analysis, Methodology, Visualization, Writing – original draft; **CK:** Investigation, Data curation, Writing – review & editing; **LT:** Resources, Supervision; **DP:** Resources, Formal analysis, Writing – review & editing, Supervision; **JW:** Conceptualization, Methodology, Software, Formal analysis, Writing – review & editing, Supervision.

