## Supplementary Material for "Effects of Subthalamic Deep Brain Stimulation on Emotion Processing in Parkinson’s Disease"

Supplementary Methods

**Exclusion criteria**

Participants were excluded if they had any other neurological or psychiatric conditions (e.g., epilepsy, other movement disorders), severe depression as indicated by the Beck’s Depression Inventory II (BDI-II > 29), cognitive impairment (Montreal Cognitive Assessment (MoCA) < 24) or significant visual or hearing impairment affecting computer-based tasks.

**Recruitment and follow-up exclusions**

Initially, 22 PwPD scheduled for bilateral DBS surgery, 24 non-DBS PwPD and 32 healthy controls completed baseline assessments. Of the 22 DBS candidates, four did not undergo surgery, and two were excluded due to meeting the exclusion criteria, resulting in 16 DBS participants (3 women), who completed follow-up assessments after undergoing STN-DBS surgery. In the non-DBS group, 20 of 24 participants completed follow-up assessments; however, four participants were subsequently excluded based on exclusion criteria, resulting in 16 participants (4 women) included in the analysis. In addition to age and sex, the PwPD groups were matched for disease duration and stage according to the Hoehn and Yahr scale (Hoehn & Yahr, 1967).

Of the initial 32 healthy controls, ten were excluded: one due to missing follow-up and nine to ensure group matching for age and sex (Figure 1).

Supplementary Tables

***Supplementary Table 1: Questionnaires and Pen-and-Paper test outcomes***

|  | **DBS** | | | **Non-DBS** | | | **HC** | | |
| --- | --- | --- | --- | --- | --- | --- | --- | --- | --- |
| Time point | BL  Mean (SD) | FU  Mean (SD) | FL - BU  Delta (p) | BL  Mean (SD) | FU  Mean (SD) | FL - BU  Delta (p) | BL  Mean (SD) | FU Mean (SD) | FL - BU  Delta (p) |
| **MoCA** | 27.88 | 27.88 | 0.00   (1.00) | 28.69 | 28.38 | -0.31 (0.37) | 28.82 | 29.18 | 0.36  (0.23) |
| **FAB** | 17.69 | 17.31 | -0.38 (0.16) | 17.56 | 17.88 | 0.31  (0.02) | 17.91 | 17.86 | -0.05 (0.58) |
| **BDI-II** | 7.69 | 6.94 | -0.75 (0.55) | 10.0 | 9.44 | -0.56 (0.57) | 3.32 | 3.82 | 0.50  (0.53) |
| **AES** | 28.69 | 31.5 | 2.81  (0.08) | 29.56 | 32.06 | 2.5  (0.20) | 25.41 | 25.59 | 0.18 (0.84) |
| **HCL-32** | 11.06 | 13.38 | 2.31   (0.11) | 12.31 | 10.69 | -1.63 (0.34) | 15.09 | 14.32 | -0.77 (0.51) |
| **QUIP-RS** | 11.13 | 12.13 | 1.00   (0.74) | 15.44 | 17.44 | 2.00  (0.28) | - | - | - |
| **PDQ-39** | 24.60 | 20.83 | -3.77 (0.13) | 20.03 | 21.67 | 1.64   (0.38) | - | - | - |

***Supplementary Table 2: Primary outcomes of emotion recognition task (Mean and SD) for all emotions by group, condition and time point***

| **Emotion** | **Group** | **Time point** | **N** | **Mean correct** | **SD** |
| --- | --- | --- | --- | --- | --- |
| **Afraid** | **DBS** | BL | 384 | 0.333 | 0.472 |
|  |  | FU | 384 | 0.279 | 0.449 |
|  | **Non-DBS** | BL | 384 | 0.383 | 0.487 |
|  |  | FU | 384 | 0.26 | 0.439 |
|  | **HC** | BL | 528 | 0.523 | 0.5 |
|  |  | FU | 528 | 0.405 | 0.491 |
| **Angry** | **DBS** | BL | 320 | 0.903 | 0.296 |
|  |  | FU | 320 | 0.916 | 0.278 |
|  | **Non-DBS** | BL | 320 | 0.8 | 0.401 |
|  |  | FU | 320 | 0.881 | 0.324 |
|  | **HC** | BL | 440 | 0.911 | 0.285 |
|  |  | FU | 440 | 0.939 | 0.24 |
| **Disgusted** | **DBS** | BL | 352 | 0.574 | 0.495 |
|  |  | FU | 352 | 0.651 | 0.477 |
|  | **Non-DBS** | BL | 352 | 0.642 | 0.48 |
|  |  | FU | 352 | 0.676 | 0.469 |
|  | **HC** | BL | 484 | 0.711 | 0.454 |
|  |  | FU | 484 | 0.756 | 0.43 |
| **Happy** | **DBS** | BL | 352 | 0.943 | 0.232 |
|  |  | FU | 352 | 0.98 | 0.14 |
|  | **Non-DBS** | BL | 352 | 0.946 | 0.226 |
|  |  | FU | 352 | 0.966 | 0.182 |
|  | **HC** | BL | 484 | 0.957 | 0.204 |
|  |  | FU | 484 | 0.973 | 0.162 |
| **Neutral** | **DBS** | BL | 352 | 0.901 | 0.3 |
|  |  | FU | 352 | 0.872 | 0.334 |
|  | **Non-DBS** | BL | 352 | 0.906 | 0.292 |
|  |  | FU | 352 | 0.938 | 0.242 |
|  | **HC** | BL | 484 | 0.897 | 0.305 |
|  |  | FU | 484 | 0.905 | 0.294 |
| **Sad** | **DBS** | BL | 352 | 0.668 | 0.472 |
|  |  | FU | 352 | 0.602 | 0.49 |
|  | **Non-DBS** | BL | 352 | 0.619 | 0.486 |
|  |  | FU | 352 | 0.585 | 0.493 |
|  | **HC** | BL | 484 | 0.694 | 0.461 |
|  |  | FU | 484 | 0.607 | 0.489 |
| **Surprised** | **DBS** | BL | 352 | 0.906 | 0.292 |
|  |  | FU | 352 | 0.884 | 0.321 |
|  | **Non-DBS** | BL | 352 | 0.892 | 0.311 |
|  |  | FU | 352 | 0.852 | 0.355 |
|  | **HC** | BL | 484 | 0.89 | 0.313 |
|  |  | FU | 484 | 0.793 | 0.405 |

***Supplementary Table 3: Two-way interaction effects of group and time point for each emotion across all groups***

| **Emotion** | **F value** | **p-value** |
| --- | --- | --- |
| **afraid** | 2.354 | 0.308 |
| **angry** | 0.262 | 0.270 |
| **disgusted** | 0.630 | 0.73 |
| **happy** | 1.354 | 0.508 |
| **neutral** | 4.174 | 0.124 |
| **sad** | 1.472 | 0.479 |
| **surprised** | 3.832 | 0.147 |

***Supplementary table 4: Primary outcomes of emotion discrimination task with mean and SD standard deviations of correct fraction per group, condition and time point***

| **Condition** | **Group** | **Time point** | **N** | **Accuracy**  **mean (sd)** | **Response time in ms**  **mean (sd)** |
| --- | --- | --- | --- | --- | --- |
| Emotional condition | DBS | BL | 959 | 0.680 (0.467) | 2832 (903) |
|  |  | FU | 958 | 0.675 (0.468) | 2977 (994) |
|  | non-DBS | BL | 960 | 0.752 (0.432) | 3001 (968) |
|  |  | FU | 958 | 0.719 (0.450) | 3039 (981) |
|  | HC | BL | 1319 | 0.790 (0.407) | 2764 (1000) |
|  |  | FU | 1320 | 0.798 (0.401) | 2732 (961) |
| Non-emotional condition | DBS | BL | 669 | 0.895 (0.306) | 1994 (767) |
|  |  | FU | 667 | 0.894 (0.309) | 2029 (885) |
|  | non-DBS | BL | 664 | 0.929 (0.257) | 1880 (836) |
|  |  | FU | 668 | 0.886 (0.318) | 2109 (947) |
|  | HC | BL | 914 | 0.951 (0.216) | 1790 (762) |
|  |  | FU | 921 | 0.951 (0.216) | 1795 (760) |

***Supplementary table 5: Go/No-Go task: impulsivity outcomes across conditions and groups***

|  | **Condition** | **Group** | **Time point** | **N** | **Accuracy**   **mean (sd)** | **Response time in ms**  **mean (sd)** |
| --- | --- | --- | --- | --- | --- | --- |
| No-Go trials | Emotional | DBS | BL | 467 | 0.938 (0.242) | *NA* |
|  |  |  | FU | 471 | 9968 (0.176) |  |
|  |  | Non-DBS | BL | 475 | 0.964 (0.186) |  |
|  |  |  | FU | 473 | 0.958 (0.201) |  |
|  |  | HC | BL | 652 | 0.969 (0.173) |  |
|  |  |  | FU | 654 | 0.965 (0.184) |  |
|  | Non-emotional | DBS | BL | 467 | 0.923 (0.267) | *NA* |
|  |  |  | FU | 472 | 0.932 (0.252) |  |
|  |  | Non-DBS | BL | 473 | 0.962 (0.192) |  |
|  |  |  | FU | 477 | 0.964 (0.186) |  |
|  |  | HC | BL | 657 | 0.974 (0.159) |  |
|  |  |  | FU | 655 | 0.983 (0.129) |  |
| Go trials | Emotional | DBS | BL | 1100 | 0.955 (0.206) | 608 (131) |
|  |  |  | FU | 1096 | 0.951 (0.217) | 626 (130) |
|  |  | Non-DBS | BL | 1101 | 0.956 (0.204) | 616 (138) |
|  |  |  | FU | 1099 | 0.944 (0.229) | 619 (132) |
|  |  | HC | BL | 1523 | 0.953 (0.212) | 602 (128) |
|  |  |  | FU | 1521 | 0.955 (0.207) | 608 (132) |
|  | Non-emotional | DBS | BL | 1089 | 0.982 (0.134) | 560 (156) |
|  |  |  | FU | 1110 | 0.965 (0.184) | 567 (153) |
|  |  | Non-DBS | BL | 1089 | 0.982 (0.134) | 550 (143) |
|  |  |  | FU | 1105 | 0.974 (0.160) | 565 (152) |
|  |  | HC | BL | 1515 | 0.991 (0.096) | 522 (139) |
|  |  |  | FU | 1519 | 0.995 (0.072) | 504 (115) |

Supplementary Figures

  
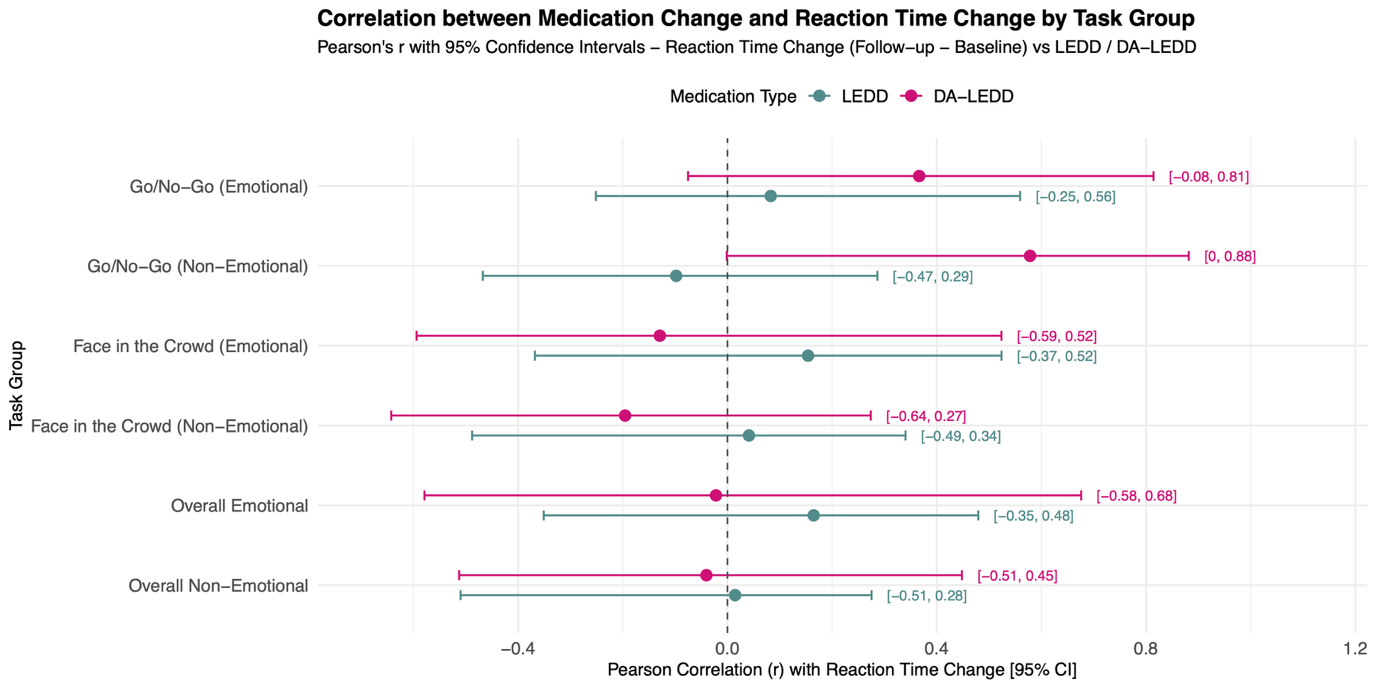

**Supplementary Figure 1:** **Correlation between medication change and reaction time change by condition by medication type (LEDD and DA-LEDD):** Horizontal bars reflect 95% confidence intervals based on 1000 bootstrap samples. Each dot represents the strength and direction of the Pearson correlation coefficient (r) between absolute change in reaction time change from baseline to follow-up and the change in medication dosage. Correlations are shown separately for total LEDD (blue) and dopamine agonist LEDD (pink). A vertical dashed line at r = 0 indicates no association. In the “overall” row, the mean reaction time across all trials per participant was used for correlation with medication change.
